# Gender differences in housework and childcare among Japanese workers during the COVID-19 pandemic

**DOI:** 10.1101/2021.07.29.21261306

**Authors:** Toshihide Sakuragi, Rie Tanaka, Mayumi Tsuji, Seiichiro Tateishi, Ayako Hino, Akira Ogami, Masako Nagata, Shinya Matsuda, Yoshihisa Fujino, CORoNaWork Project

## Abstract

**Objectives:** Although gender stereotypes regarding paid work and unpaid work are changing, most wives are responsible for taking care of the family and home in Japan. It is unclear how time spent on housework and childcare has changed between working men and women during the COVID-19 pandemic in Japan. The purpose of this study is to investigate how working men and women’s responsibilities for housework and childcare changed during the COVID-19 pandemic in Japan depending on occupation, job type, and the number of employees in the workplace.

**Methods:** A cross-sectional analysis (N=14,454) was conducted using data from an internet monitoring study (CORoNa Work Project), which was conducted in December 2020. A multilevel logistic model with nested prefectures of residence was conducted to estimate the odds ratio (OR) for change in time devoted to housework and childcare among men and women adjusting for age, household income, frequency of telecommuting, presence of spouse who work, occupation, job type, the number of employees in the workplace, and the incidence rate of COVID-19 by prefecture.

**Results:** Regardless of occupation related factors, more women than men reported increased time spent on housework and childcare. Furthermore, women were significantly more likely to experience an increase in time spent on those activities than men (housework: OR 1.97, 95% CI [1.75, 2.21], *p* < 0.001; childcare: OR 1.66, 95% CI [1.37, 2.02], *p* < 0.001).

**Conclusions:** The time spent by women on housework and childcare increased significantly compared to men during the COVID-19 pandemic in Japan.

## Introduction

Women tend to spend more time on housework than men. This is true in Japan, where the gender difference is greater than in other countries.^1^ Time spent on housework and childcare by husbands and wives with a child or children under six years old is 1.23 and 7.34 hours per day, respectively.^1^ Regarding hours spent exclusively on childcare per day, husbands spend 0.49 hours and women spend 3.45 hours each day. ^1^

In Japan, the participation of women in the labor force has increased in recent years; the employment rate of Japanese women in the working age population was 69.6% in 2018, which ranked 16^th^ among 35 OECD countries.^2^ According to the White Paper on Gender Equality 2020, for women aged 25–29, time devoted to housework, childcare, and caregiving has remarkably decreased over the past three decades, while work-related time has increased.^3^

In terms of work allocation between couples, husbands, especially men in their child-rearing years, overwhelmingly work longer hours than their wives.^3^ In a 2009 study examining employment and household tasks among Japanese couples, women who were employed full-time spent 21 hours a week on housework, whereas their husbands only spent 5 hours a week on such work.^4^

Multiple Japanese laws, such as the Childcare Leave Law and the Female Success Promotion Law, have been enforced since the 1990s with the aim of improving the balance between work and family, and of promoting gender equality. However, gender disparities still exist. For example, 83% of women utilize childcare leave compared to 5% of men.^5,6^ In addition, childcare leave laws vary by industry and business size. More than 90% of businesses in the information technology and education sectors have childcare leave, whereas only 60% of businesses in the construction and manufacturing sectors have it. Childcare leave has been introduced in 99% of organizations with more than 500 employees but only in 71% of organizations with fewer than 30 employees.^7^

During the COVID-19 pandemic, there has been a change in time spent on housework and childcare, which has been the subject of studies in several countries.^8-10^ In a UK study examining the impact of the COVID-19 lockdown on unpaid care work, researchers found that women spent significantly more time on housework and childcare duties than men, resulting in higher levels of psychological distress in women.^11^ However, there are very few similar studies in Japan, where women are responsible for more unpaid workhours than men.^12^ In Japan, women significantly increased their time performing housework after the start of the COVID-19 pandemic in Tokyo and three surrounding prefectures.^13^ In the present study, we investigate gender differences in changes in time spent on housework and childcare after the start of the COVID-19 pandemic throughout Japan, considering occupation related factors, including frequency of telecommuting, occupation, job type, and the number of employees in the workplace.

## Methods

### Study design and participants

This study used data from the Collaborative Online Research on the Novel-coronavirus and Work (CORoNa Work) Project, which was an Internet-based prospective cohort study of workers assigned by prefecture, job type, and gender conducted during December 2020 in Japan. The survey comprised of workers aged 20–65 years, and additional details of the CORoNa Work Project have been reported previously.^14^ The protocol for the CORoNa Work Project was approved by the Ethics Committee of the University of Occupational and Environmental Health, Japan. The CORoNa Work Project collected data from 33,302 individuals. After excluding invalid responses, 27,036 individuals were included in the final data set. In our study, 14,454 individuals who live with their spouse were included. In addition, 4,189 individuals who reported the presence of pre-school and/or elementary school children in the home were included in our analyses regarding childcare (Figure 1).

**Figure 1.**
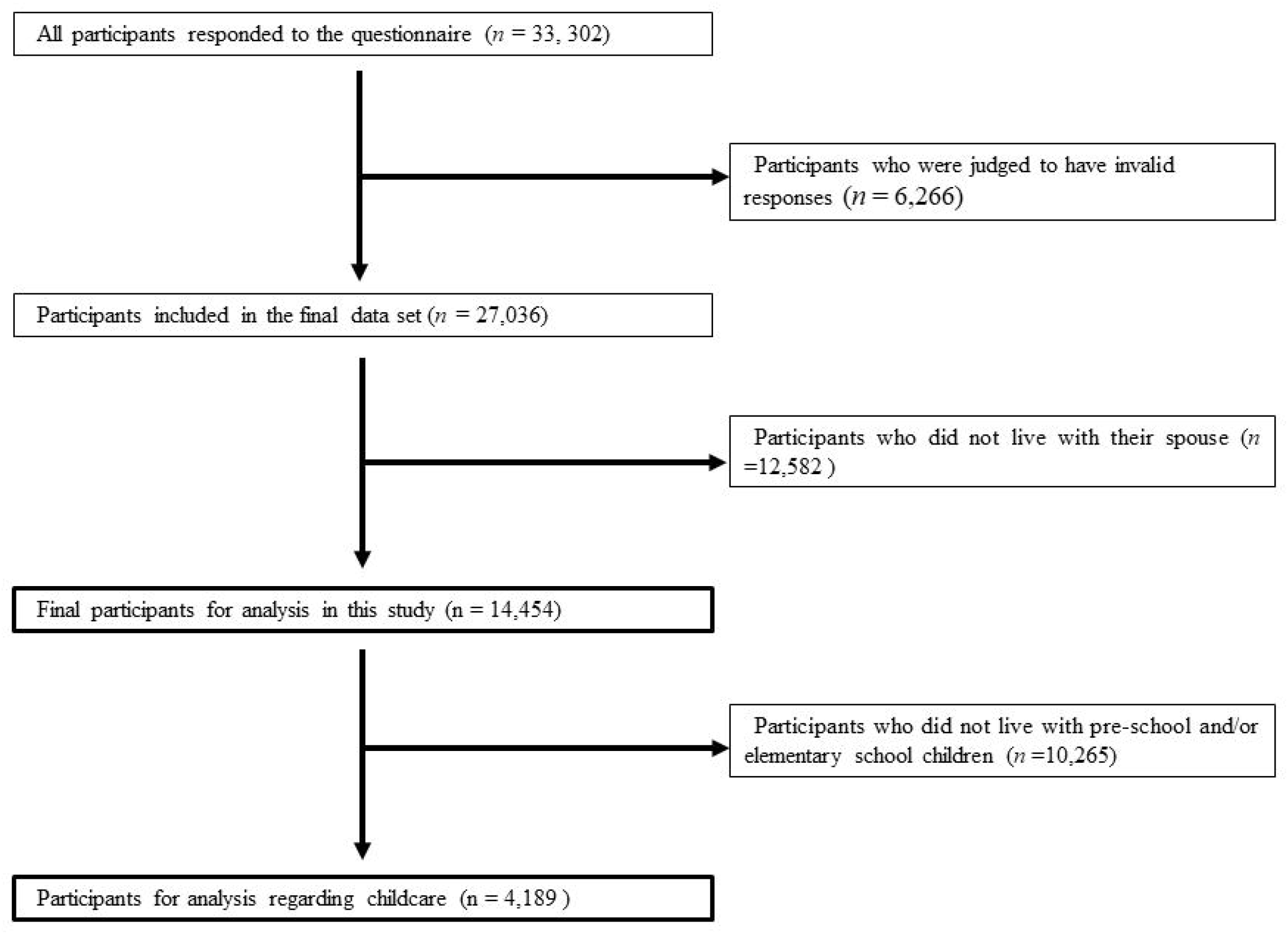
Flowchart for the inclusion of this study participants

### Work-related factors

Occupations were categorized into the following seven groups: company worker (general employee, manager, executive manager); public employee, faculty member, and non-profit organization employee; temporary/contract employee; self-employed (commercial and industrial services, small office/home office); agriculture, forestry, and fishing; professional occupation (lawyer, tax accountant, medical-related, etc.); and other occupation.

Job type was assessed with a question requiring the respondent to select the closest job description. Answer categories were as follows: mainly desk work (e.g., office work, computer work); mainly work involving talking with people (e.g., customer service, sales, etc.), and manual work (e.g., work at a production site, manual labor, nursing care, etc.).

Respondents provided the number of employees in their workplace, by selecting one of the following numerical ranges: 1 person (freelance), 2–4, 5–9, 10–29, 30–49, 50–99, 100–499, 500–999, 1,000–9,999, 10,000 or more. In our study, responses were divided as follows: 1–29, 30–99, 100–999, and >1000.

Frequency of telecommuting were categorized into the following five groups: 4 days a week or more, 2 days a week or more, more than 1 day a week, more than 1 day a month, almost never.

### Housework and Childcare

The survey addressed the impact of the COVID-19 pandemic with the following set-up explanation: “We would like to ask you about the impact of the outbreak of COVID-19 on your social and living conditions.” Then, particular issues were addressed underneath. For “Time spent on housework,” the following answer options were presented: increased, stayed the same, and decreased. Similarly, for “Time spent on childcare,” the same answer options were presented: increased, stayed the same, and decreased.

### Other variables

Information on age (continuous), presence of pre-school and/or elementary school children (absence or presence), presence of spouse who work, and household income were also provided by survey respondents. In our study, household income (million yen) was categorized into the four following groups: 0.5–2.49, 2.50–3.74, 3.75–4.89, and 4.90 and higher.

### Statistical analysis

This study evaluates changes in time spent on housework/childcare by gender and work-related factors. After excluding respondents who reported a decrease in time spent on these activities (approximately 3% of respondents), we examined the association between increased time spent on housework/childcare and gender. Multilevel logistic regression analysis was performed using the following factors: prefecture of residence, age, household income, occupation, job type, the number of employees in the workplace, and the incidence rate of COVID-19 by prefecture. Only respondents with pre-school and/or elementary school children were included in the childcare analysis. Statistical analyses were conducted using Stata/IC 14.0 (StataCorp, College Station, TX, USA). Statistical significance was set at *p* < .05.

## Results

Table 1 shows the respondents’ characteristics. The mean age of respondents was 49.3±9.9 years old. The percentage of respondents who reported their household income as 0.5–2.49, 2.50–3.74, 3.75–4.89, and 4.90 and higher was 17.5%, 24.0%, 28.1%, and 30.4%, respectively. More than half of respondents were company workers (60.7%). Public employee/faculty member/non-profit organization employees, temporary/contract employees, self-employed employees, professional occupations, and other occupations were 12.7%, 8.2%, 9.3%, 6.8%, and 1.7%, respectively. According to job type, about half of respondents were engaged in desk work (51.4%). Respondents engaged in mainly talking to others or manual work were 25.4% and 23.2%, respectively. In terms of the number of employees in the workplace, 31.3%, 14.5%, 27.2%, and 27.1% of respondents worked in workplaces with 1–29, 30–99, 100–999, and >1000 employees, respectively. Regarding telecommuting, 78.2% of respondents reported they almost never telecommute. Respondents with the presence of spouse who work were 74.8%.

**Table 1.**
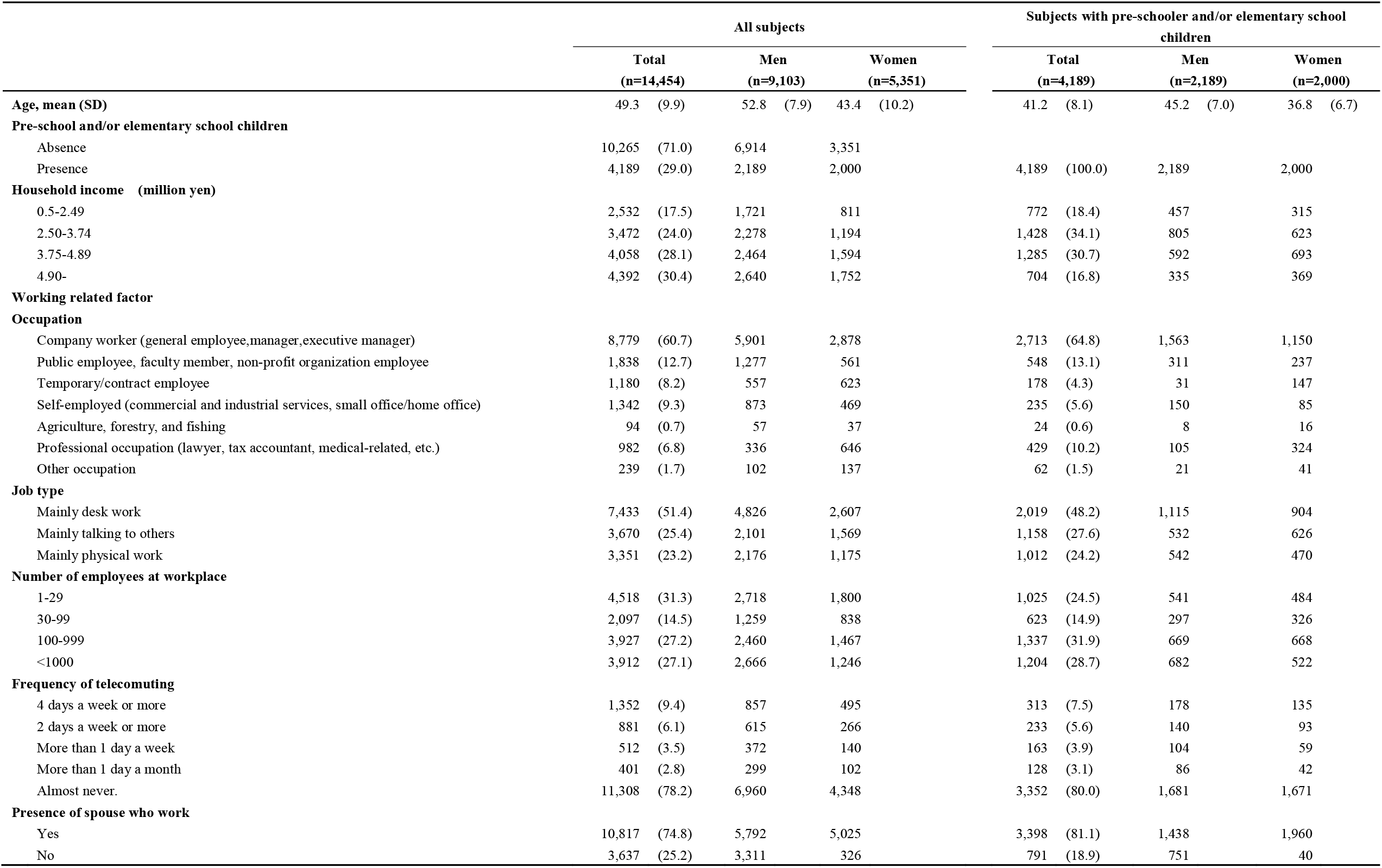
Characteristics of the subjects.

Figure 2a and 2b display the changes in housework during the pandemic by work-related factors and gender. Regardless of occupation, a greater percentage of women reported increased time for housework compared with men. Regardless of frequency of telecommuting, job type or the number of employees in the workplace, a greater percentage of women reported increased time for housework compared with men.

**Figure 2.**
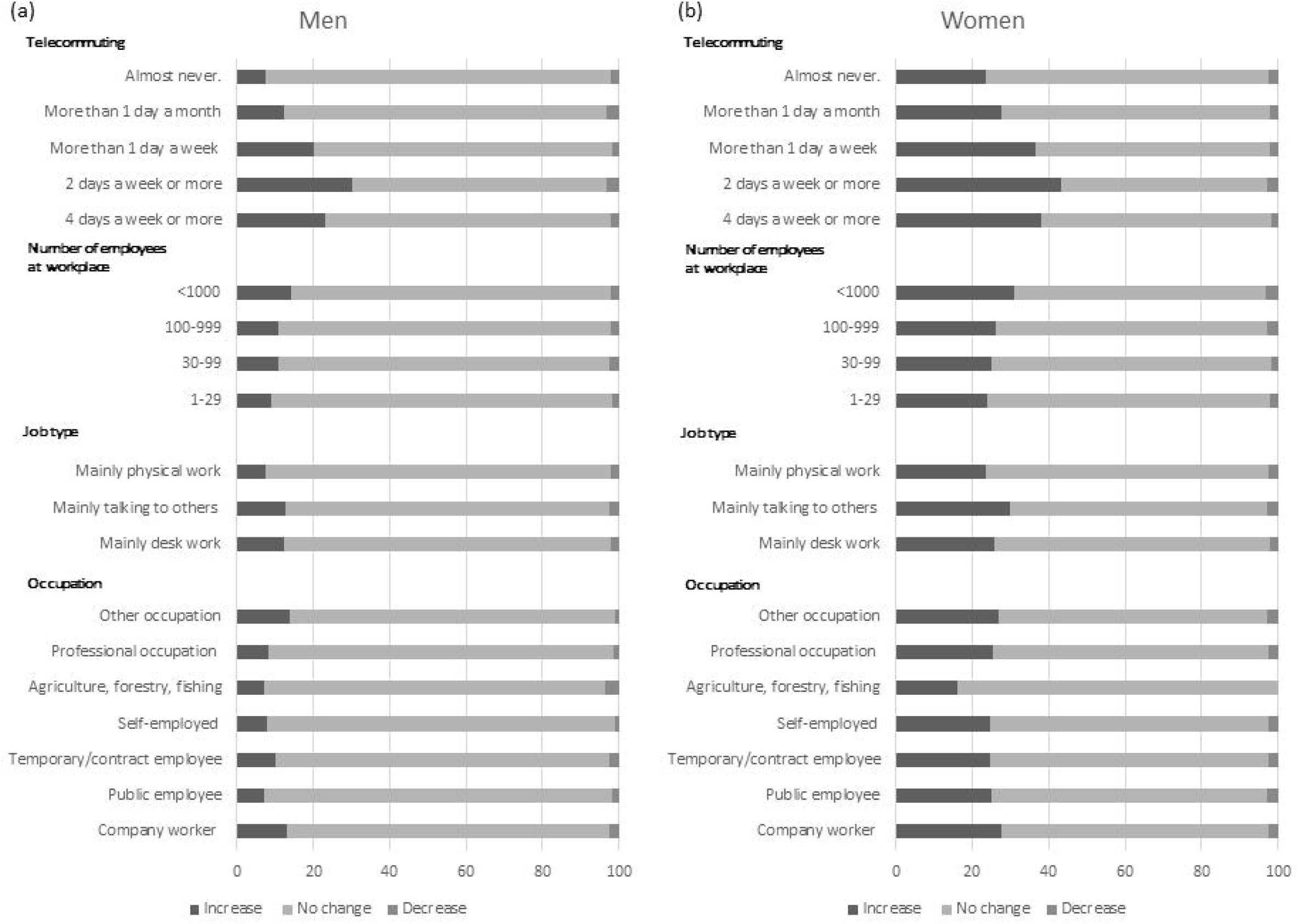
Change in time spent on housework during the COVID-19

Figure 3a and 3b display the changes in childcare during the pandemic by work-related factors and gender among respondents with pre-school and/or elementary school children. Regardless of occupation, a greater percentage of women reported increased time for childcare compared with men. Regardless of frequency of telecommuting, job type or the number of employees in the workplace, a greater percentage of women reported increased time for childcare compared with men.

**Figure 3.**
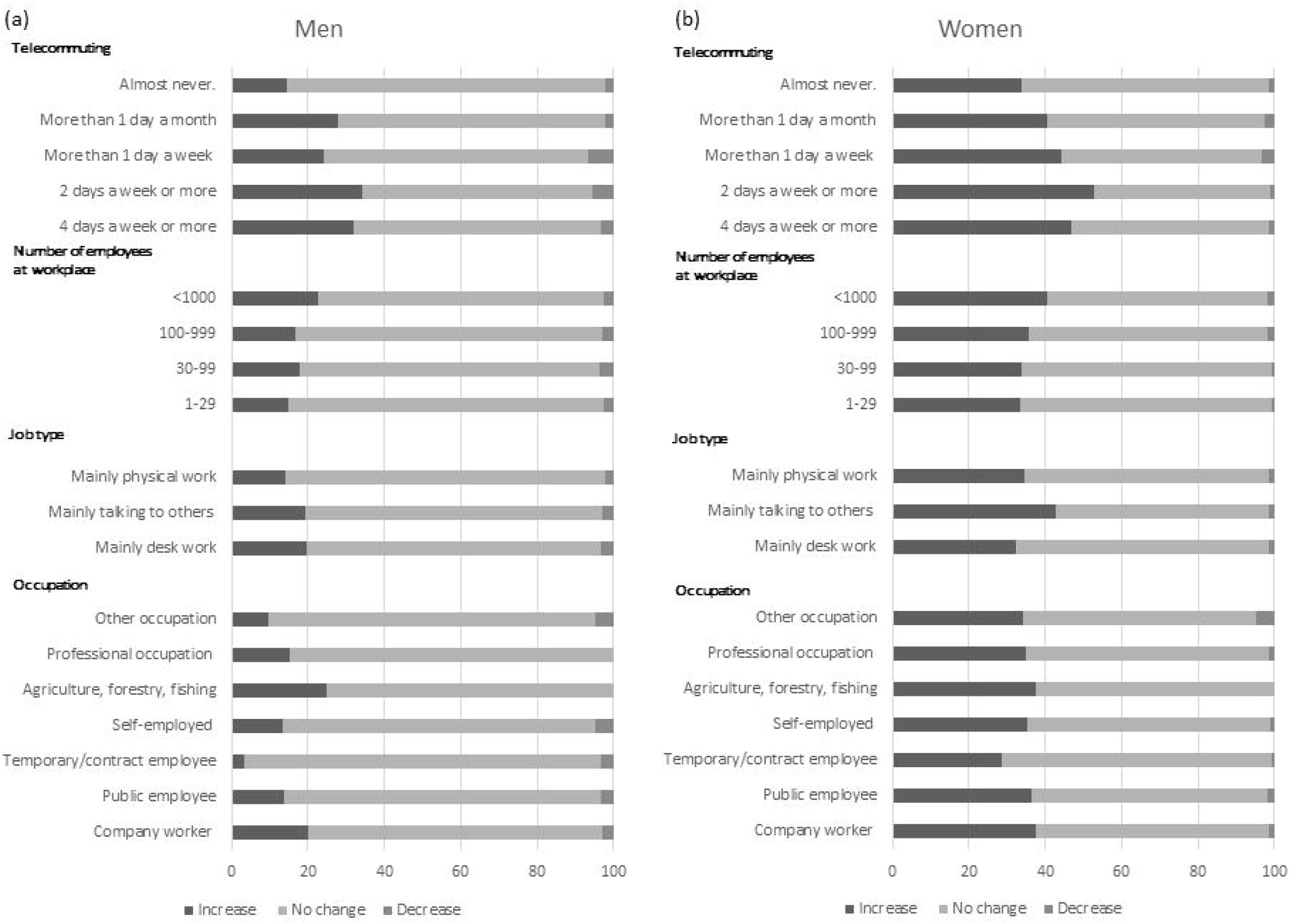
Change in time spent on childcare during the COVID-19

Table 2 provides the results of the multilevel logistic regression analysis regarding associations between gender and change in time spent on housework during the pandemic. Women were significantly more likely than men to report increased time for housework, after adjusting for age (OR 1.91, 95% CI [1.73, 2.12]). After adjusting for age, household income, frequency of telecommuting, presence of spouse who work, occupation, job type, the number of employees in the workplace, and the incidence rate of COVID-19 by prefecture, the association remained significant (OR 1.97; 95% CI [1.75, 2.21]). Similarly, table 3 provides the results of the multilevel logistic regression analysis regarding associations between gender and change in time spent on childcare during the pandemic. Women were significantly more likely than men to report increased time for childcare, after adjusting for age (OR 1.51, 95% CI [1.27, 1.79]). After adjusting for age, household income, frequency of telecommuting, presence of spouse who work, occupation, job type, the number of employees in the workplace, and the incidence rate of COVID-19 by prefecture, the association remained significant (OR 1.66, 95% CI [1.37, 2.02]).

**Table 2.** Results of multivariable analysis in the relationship between sex and change of the time spent on housework during normal times vs during COVID-19 epidemic time.

|  | No change<br>(n=11,693 ) | Increase<br>(n=2,434) | age adjusted |  | multivariate* |  |
| --- | --- | --- | --- | --- | --- | --- |
|  |  |  | OR (95%CI) | P Value | OR (95%CI) | P Value |
| Men | 7,881 | 1,024 | 1.00 (Ref) |  | 1.00 (Ref) |  |
| Women | 3,812 | 1,410 | 1.91 (1.73-2.12) | <0.001 | 1.97 (1.75-2.21) | <0.001 |

\*:The odds ratios (OR) were estimated by a multilevel logistic model nested in the prefecture of residence, adjusting for age, household income, frequency of telecommuting, presence of spouse who work, occupation, job type, number of employees at workplace and the incidence rate of COVID-19 by prefecture.

**Table 3.** Results of multivariable analysis in the relationship between sex and change of the time spent on childcare during normal times vs during COVID-19 epidemic time.

|  |  |  | age adjusted |  | multivariate* |  |
| --- | --- | --- | --- | --- | --- | --- |
|  | No change<br>(n=2,977) | Increase<br>(n=1,122) | OR (95%CI) | P<br>Value | OR (95%CI) | P Value |
| Men | 1,728 | 399 | 1.00 (Ref) |  | 1.00 (Ref) |  |
| Women | 1,249 | 723 | 1.51 (1.27-1.79) | <0.001 | 1.66 (1.37-2.02) | <0.001 |

\*:The odds ratios (OR) were estimated by a multilevel logistic model nested in the prefecture of residence, adjusting for age, household income, frequency of telecommuting, presence of spouse who work, occupation, job type, number of employees at workplace and the incidence rate of COVID-19 by prefecture.

## Discussion and Conclusion

This study demonstrated that during the COVID-19 pandemic, the percentage of women who perceived that they spent more time on housework and childcare was significantly greater than that of men after adjusting for occupation-related factors, including frequency of telecommuting, occupation, job type, and the number of employees in the workplace. One possible reason for this increase in women’s housework is that men were unable to adapt to domestic work during the spread of COVID-19. Even before the pandemic, there was a difference in the time men and women spent on housework and childcare.^15^ Although the participation rate of men in housework is increasing each year, a 2009 report found that husbands of women who work part-time or full-time perform only 11% or 18%, respectively, of housework duties. ^4^ Women with young children spend more time doing housework than women without young children, but men spend the same amount of time doing housework regardless of having children.^4^ Compared to Western countries, Japanese men spend less time on housework.^7^ For example, in Japan, wives spend an average of 263 minutes on housework wives, whereas husbands spend 37 minutes.^16^ Awareness of stereotypes regarding gender roles has increased in Japan; the percentage of men and women who disagree with the idea that “husbands should work outside and wives should take care of their home” has increased in recent years.^3^ However, there is no change regarding “work-related time” spent by husbands; many men work 60 hours or more per week. Thus, there seems to be a gap between awareness and reality in Japan. Although men might want to participate more in housework, it is possible they do not know how to increase their participation, especially during the COVID-19 pandemic.

For women, employment changes during the pandemic may be linked to our findings. Globally, the COVID□19 pandemic has caused an increase in unemployment.^17^ In particular, women’s employment patterns have changed dramatically. In the Netherlands, inequality between men and women regarding paid work has widened during the pandemic compared to before the lockdown.^9^ Yanfei et al. reported that the average working hours of female workers have decreased more than that of male workers during the COVID-19 pandemic in Japan.^18^ Jobs involving face-to-face contact, such as those in the food service and retail industries, have been particularly affected by the COVID-19 pandemic. Our survey also found that women whose jobs involve mainly talking to others had a larger increase in time spent on housework and childcare than women whose jobs involve mainly desk work and mainly manual work. Thus, the increase may be because women spent more time at home during the pandemic due to lob loss or a reduction in work hours.

Focusing exclusively on childcare, school closures throughout Japan impacted working mothers. When elementary schools and kindergartens closed due to COVID-19, children stayed at home. In Japan, the nuclear family has become the predominant family model,^11^ and the number of dual-income families has increased 1.7 times in the past 30 years.^19^ For working mothers who usually rely on their parents for childcare, that childcare option was eliminated because people were forced to avoid contact with people except for family members living together to prevent the spread of COVID-19. As a result, many women with young children changed the way they worked, including in terms of unpaid work. Consequently, during the pandemic, mothers tended to spend more time on childcare than before. In contrast to the increase in housework time, an increase in childcare time may have benefits; increased time spent on childcare is associated with good health, whereas increased time spent on housework is associated with poor health, particularly among women.^20^

The Japanese government is accelerating efforts to encourage childcare leave.^21^ However, the type of job and the size of the company may impact whether it is easy to take leave. ^22^ In our study, we also found a larger increase in time spent on housework and childcare for men and women working for companies with a large number of employees. Those who work in a workplace where it is easier to take childcare leave during normal times may be more likely to take childcare leave in emergency times, such as the COVID-19 pandemic.

This study has several limitations. First, this study used a data set obtained from an online survey. Considering that the survey respondents were individuals who had access to the Internet and were interested in the survey, the results cannot be generalized. ^23^ Second, this study did not examine changes in the actual amount of time spent on housework and childcare because the questionnaire did not request that information. Third, the data did not contain detailed information about the respondents’ home situations. For example, it may be useful to examine the impact availability of close relatives, other family background information, and school status on changes in housework and childcare. Fourth, the survey did not obtain information about the employment status of a partner, which may impact housework and childcare.

In conclusion, regardless of occupation, job type, and the number of employees in the workplace, the time spent by women on housework and childcare increased significantly compared to men, suggesting that the burden on women may be increasing during the COVID-19 pandemic.

## Data Availability

Data not available due to ethical restrictions

## Acknowledgements

This study was supported and partly funded by the research grant from the University of Occupational and Environmental Health, Japan (no grant number); Japanese Ministry of Health, Labour and Welfare (H30-josei-ippan-002, H30-roudou-ippan-007, 19JA1004, 20JA1006, 210301-1, and 20HB1004); Anshin Zaidan (no grant number), the Collabo-Health Study Group (no grant number), and Hitachi Systems, Ltd. (no grant number) and scholarship donations from Chugai Pharmaceutical Co., Ltd. (no grant number)

The current members of the CORoNaWork Project, in alphabetical order, are as follows: Dr. Yoshihisa Fujino (present chairperson of the study group), Dr. Akira Ogami, Dr. Arisa Harada, Dr. Ayako Hino, Dr. Hajime Ando, Dr. Hisashi Eguchi, Dr. Kazunori Ikegami, Dr. Kei Tokutsu, Dr. Keiji Muramatsu, Dr. Koji Mori, Dr. Kosuke Mafune, Dr. Kyoko Kitagawa, Dr. Masako Nagata, Dr. Mayumi Tsuji, Ms. Ning Liu, Dr. Rie Tanaka, Dr. Ryutaro Matsugaki, Dr. Seiichiro Tateishi, Dr. Shinya Matsuda, Dr. Tomohiro Ishimaru, and Dr. Tomohisa Nagata. All members are affiliated with the University of Occupational and Environmental Health, Japan.

## Disclosure

### Ethical approval

This study was approved by the ethics committee of the University of Occupational and Environmental Health, Japan (reference No. R2-079 and R3-006).

### Informed Consent

Informed consent was obtained in the form of the website.

### Registry and the Registration No. of the study/Trial

N/A

### Animal Studies

N/A

## References

1. Office GEBC. Women and Men in Japan 2020. Work-life Balance 2020; https://www.gender.go.jp/english_contents/pr_act/pub/pamphlet/women-and-men20/index.html https://www.gender.go.jp/english_contents/pr_act/pub/pamphlet/women-and-men20/pdf/1-4.pdf. Accessed July, 15, 2021.

2. Office GEBC. Women and Men in Japan 2020. Work 2020; https://www.gender.go.jp/english_contents/pr_act/pub/pamphlet/women-and-men20/index.html https://www.gender.go.jp/english_contents/pr_act/pub/pamphlet/women-and-men20/pdf/1-3.pdf. Accessed July, 15, 2021.

3. Office GEBC. White Paper. Balancing “Work” and “Housework/Childcare/Caregiving” - How do Individuals, Households, and Society Face the Issue? -(From the “White Paper on Gender Equality 2020”) 2020; https://www.gender.go.jp/english_contents/about_danjo/whitepaper/index.html https://www.gender.go.jp/english_contents/about_danjo/whitepaper/pdf/ewp2020.pdf. Accessed July, 15, 2021.

4. Tsuya NO, Bumpass LL, Choe MK, Rindfuss RR. Employment and Household Tasks of Japanese Couples, 1994-2009. Demographic research. 2012;27:705–718.

5. Office GEBC. The situation of gender equality society in international comparison. 2003.

6. Office GEBC. 2017 Equal Employment Basic Survey in japan. 2007.

7. Office GEBC. The White paper on gender equality 2020. 2020.

8. Del Boca D, Oggero N, Profeta P, Rossi M. Women’s and men’s work, housework and childcare, before and during COVID-19. Review of economics of the household. 2020:1–17.

9. Yerkes MA, André SCH. ‘Intelligent’ lockdown, intelligent effects? Results from a survey on gender (in)equality in paid work, the division of childcare and household work, and quality of life among parents in the Netherlands during the Covid-19 lockdown. 2020;15(11):e0242249.

10. Shafer K, Scheibling C, Milkie MA. The Division of Domestic Labor before and during the COVID-19 Pandemic in Canada: Stagnation versus Shifts in Fathers’ Contributions. Canadian review of sociology = Revue canadienne de sociologie. 2020;57(4):523–549.

11. Xue B, McMunn A. Gender differences in unpaid care work and psychological distress in the UK Covid-19 lockdown. PloS one. 2021;16(3):e0247959.

12. Office GEBC. Seikatsujikannokokusaihikaku. 2021; https://www.gender.go.jp/about_danjo/whitepaper/r02/zentai/html/column/clm_01.html. Accessed July, 15, 2021.

13. Suka M, Yamauchi T, Yanagisawa H. Changes in health status, workload, and lifestyle after starting the COVID-19 pandemic: a web-based survey of Japanese men and women. Environmental health and preventive medicine. 2021;26(1):37.

14. Fujino Y, Ishimaru T, Eguchi H, et al. Protocol for a nationwide Internet-based health survey in workers during the COVID-19 pandemic in 2020. medRxiv. 2021.

15. Office GEBC. Women and Men in Japan 2018. 2018; https://www.gender.go.jp/english_contents/pr_act/pub/pamphlet/women-and-men18/index.html https://www.gender.go.jp/english_contents/pr_act/pub/pamphlet/women-and-men18/pdf/1-4.pdf. Accessed July, 20, 2021.

16. National Institute of Population and Social Security Research Tokyo J. The 6th National Survey on Family in Japan, 2018 2018.

17. Organization IL. As job losses escalate, nearly half of global workforce at risk of losing livelihoods. 2020; https://www.ilo.org/global/about-the-ilo/newsroom/news/WCMS_743036/lang--en/index.htm. Accessed July, 20, 2020.

18. Yanfei Z. How Women Bear the Brunt of COVID-19’s Damages on Work (Continued): The Gender Gap in Employment Recovery. 2021.

19. Ministry of Health LaW. Overview of the 2019 National Life Basic Survey. 2019.

20. Jonsson KR, Oberg G, Samkange-Zeeb F, Adjei NK. Determinants and impact of role-related time use allocation on self-reported health among married men and women: a cross-national comparative study. BMC public health. 2020;20(1):1204.

21. Ministry of Health LaW. Promotion of Balancing Work and Family. https://www.mhlw.go.jp/english/policy/children/work-family/index.html. Accessed July, 20, 2020.

22. Ministry of Health LaW. Promotion of Balancing Work and Family. Introduction to the revised Child Care and Family Care Leave Law https://www.mhlw.go.jp/english/policy/children/work-family/index.html https://www.mhlw.go.jp/english/policy/affairs/dl/05.pdf Accessed July, 20, 2020.

23. Andrade C. The Limitations of Online Surveys. Indian journal of psychological medicine. 2020;42(6):575–576.

